# *NOTCH3* variants are common in the general population and associated with stroke and vascular dementia: an analysis of 200,000 participants

**DOI:** 10.1101/2020.12.14.20248151

**Authors:** Bernard PH Cho, Stefania Nannoni, Eric L Harshfield, Daniel J Tozer, Stefan Gräf, Steven Bell, Hugh S Markus

## Abstract

**Background:** Cysteine-altering *NOTCH3* variants identical to those causing the rare monogenic form of stroke, CADASIL, have been reported more common than expected in the general population, but their clinical significance and contribution to stroke and dementia risk in the community remains unclear.

**Methods:** Cysteine-altering *NOTCH3* variants were identified in UK Biobank whole-exome sequencing data (N=200,632). Frequency of stroke, dementia and other clinical features of CADASIL, and MRI white matter hyperintensity volume were compared between variant carriers and non-carriers. MRIs from those with variants were visually rated, each matched with three controls.

**Results:** Of 200,632 participants with exome sequencing data available, 443 (∼1 in 450) carried 67 different cysteine-altering *NOTCH3* variants. After adjusting for age, sex, and ancestry principal components, *NOTCH3* variant carriers had increased risk of stroke (OR: 2.33, p=0.0003), and vascular dementia (OR: 5.03, p=0.007), and increased WMH volume (standardised difference: 0.52, p<0.001), and white matter ultrastructural damage on DTI-PSMD (standardised difference: 0.71, p<0.001). On visual analysis of MRIs from 47 carriers and 148 matched controls, variants were associated with presence of lacunes (OR: 4.83, p<0.001) and cerebral microbleeds (OR: 3.61, p<0.001). WMH prevalence was most increased in the anterior temporal lobes (OR: 6.92, p<0.001) and external capsule (OR: 12.44, p<0.001).

**Conclusions:** Cysteine-changing *NOTCH3* variants are common in the general population and are risk factors for apparently “sporadic” stroke and vascular dementia. They are associated with MRI changes of SVD, in a distribution similar to that seen in CADASIL.

## INTRODUCTION

Cerebral small vessel disease (SVD) is a major cause of stroke and dementia.[1,2] Most cases are sporadic, although polygenic risk factors appear important.[3] SVD is the most common type of ischaemic stroke caused by single-gene disorders; the most common is cerebral autosomal dominant arteriopathy with subcortical infarcts and leukoencephalopathy (CADASIL), which results in early onset lacunar stroke and dementia, and is caused by distinctive *NOTCH3* variants that lead to an extra unpaired cysteine residue in one of 34 Epidermal Growth Factor-Like (EGFR) domains.[4]

Although CADASIL is considered rare, with a reported disease prevalence of 4 per 100,000,[5,6] recently a much higher frequency of typical cysteine-altering *NOTCH3* variants, as high as 1 in 400.[7,8] These could contribute to the risk of apparently sporadic lacunar stroke, but the clinical significance of these variants remains uncertain. Studies have mostly been conducted on anonymised genome-sequencing databases,[7] and it is unclear whether such apparently ‘asymptomatic’ variants have clinical implications.[9] It has been demonstrated that such variants are more likely to be in EGFR domains 7-34, while variants within clinical CADASIL cases are more likely, although not exclusively, in domains 1-6.[10]

To determine whether these variants increase disease, analysis of sequencing databases in which clinical data is also available is required. We used whole-exome sequence data from 200,632 participants in UK Biobank to determine the frequency of *NOTCH3* variants, and whether they were associated with stroke and dementia. In addition, in the subset of individuals in whom MRI scanning was performed, we correlated presence of *NOTCH3* variants with MRI markers of SVD including white matter hyperintensities (WMH) and lacunar infarcts and determined whether variant carriers had a similar spatial distribution of lesions to that seen in typical CADASIL cases.

## METHODS

### Study population

UK Biobank is a prospective study of more than 500,000 healthy volunteers aged 40-69 years recruited across the UK between 2006-2010.[11] About 9 million individuals were invited to join, of whom 5.5% participated in the baseline assessment.[12] Extensive phenotypic data were collected through questionnaires and physical examinations. A subset of about 100,000 individuals are also taking part in the MRI Study, and approximately 40,000 brain MRIs were available at the time of this analysis. These participants were selected on the basis of travelling distance from the imaging centre and not clinical information. All MRIs were performed on one of two identical Skyra 3.0T scanners (Siemens Medical Solutions, Germany). Identical acquisition parameters and quality control was used for all scans.[13] In October 2020, whole-exome sequences of 200,632 UK Biobank participants were released and all cases, were included. This analysis was performed under UK Biobank application number 36509. UK Biobank received ethical approval from the NHS National Research Ethics Service North West (11/NW/0274).

### Ascertainment of NOTCH3 pathogenic variants

The variants located in the *NOTCH3* gene region (chromosome 19:15,159,038-15,200,995, reference genome assembly GRCh38) were extracted from the exome data in PLINK format. The extracted variants were annotated using Ensembl Variant Effect Predictor.[14] Missense variants that lead to the gain or loss of a cysteine residue in one of the 34 EGFR domains of the NOTCH3 protein (amino acid position 40-1373, Uniprot accession number: Q9UM47) were identified.

### Phenotypic data fields

History of vascular risk factors, measured blood pressure, blood pressure medication and parental history of stroke, were recorded. History of diseases, including stroke, vascular dementia, epilepsy, depression, migraine, myocardial infarction were determined from self-report, hospital and death records (code list in **Supplementary Table 1**). The Framingham cardiovascular risk score was calculated.[15] Results of cognitive tests[16] including pairs matching, reaction time, prospective memory, fluid intelligence, numeric memory, trail making test part B and the symbol digit substitution test, were extracted and converted into z-scores. The average of all z-scores was used as an estimate of global cognitive function.[17] The following blood test were extracted (cholesterol, low-density lipoprotein, high-density lipoprotein, apolipoprotein A, apolipoprotein B, lipoprotein A, triglycerides, glycated haemoglobin, glucose, C-reactive protein, creatinine, cystatin C and urea),

### Brain imaging analysis

In the 19,686 participants with MRI available, measures were compared between variant subjects and controls. UK Biobank neuroimaging working group derived MRI measures for brain volume and WMH volume were used, generated by an image-processing pipeline developed and run on behalf of UK Biobank.[13] Brain volume was estimated by SIENAX,[18] and normalised for head size. WMH were quantified on fluid attenuated inversion recovery (FLAIR) images through the brain intensity abnormality classification algorithm (BIANCA).[19] In addition, the Peak width Skeletonised Mean Diffusivity (PSMD)[20] was derived in house from diffusion tensor imaging (DTI) data and is a marker of diffuse white matter damage.[19] Additionally, visual review of MRI scans (T1-weighted, T2 FLAIR and susceptibility-weighted imaging (SWI)) from those cases with *NOTCH3* variants and MRI available (N=47) was performed by a neurologist (S.N.). Each variant case was matched with three controls for age, sex, ethnicity and family history of stroke. Scans were analysed blinded to subject identity. Lacunar infarcts, cerebral microbleeds, and the spatial distribution and extent of WMH were quantified. Lacunes were counted on FLAIR images, defined as a round or ovoid subcortical fluid-filled cavity of 3-15 mm in diameter in the territory of a single perforating arteriole.[21] Cerebral microbleeds were counted using the brain observer microbleed scale,[22] defined as round well-defined hypointense foci on SWI with a diameter of 2-10 mm. The spatial distribution and extent of WMH in different brain regions was quantified on T2 FLAIR images using the modified Schelten’s scale.[23]

### Calculation of polygenic risk score

To compare the risk conferred by *NOTCH3* variants with that by common stroke variants we calculated an ischaemic stroke polygenic risk score (PRS) using more than 3 million common genetic variants.[24] We performed linear regression of the PRS adjusting for the first 10 ancestry principal components in UK Biobank to account for known differences in PRS performance across ethnic groups.[25] Standardised residuals from this regression model were used for analyses. We calculated the odds ratio (OR) for ischaemic stroke in *NOTCH3* carriers compared to non-carriers and the OR for a 1 standard deviation (SD) increase in the ischaemic stroke PRS, dividing the former by the latter (on the log-scale, assuming a linear association) to estimate the increment of the PRS in SD that was predicted to be equivalent to the risk conferred by NOTCH3 variants.[26] To examine whether the ischaemic stroke PRS affected *NOTCH3* penetrance performed statistical tests for interaction (multiplicative and additive) between the PRS and *NOTCH3* status as well as divided our sample into three groups based on their PRS: low (bottom 20% of PRS), intermediate, and high (top 20%) risk.[27]

### Statistical analysis

The effect of *NOTCH3* variant on phenotype was assessed by linear regression for continuous outcomes and logistic regression for binary outcomes. For the former, effect sizes were standardised to enable a more informative comparison. Firth’s correction was applied to all logistic regression models to account for rare event bias.[24] All regression models were adjusted for age, sex, and the first 10 principal components of ancestry. An appropriate transformation was applied to any data fields that did not follow a normal distribution: natural log (HDL, triglycerides, CRP and WMH), inverse normalisation (Lp(a), Framingham cardiovascular risk score, and the regional WMH scores) or square root (cerebral microbleed and lacune counts). When comparing demographic and risk factor profiles between variant and non-variant groups, Pearson’s Chi-squared test was used for categorical variables and the two-sample t-test was used for continuous variables. All statistical analyses were performed using R 3.6.2 and Stata 15.1 with 2-sided p-values and p <0.05 for statistical significance.

## RESULTS

### The prevalence of *NOTCH3* pathogenic variants

Of the 200,632 participants with exome sequencing data, 443 (2.2 per 1000) carried *NOTCH3* variants leading to an unpaired cysteine residue in one of the 34 EGFR domains of the NOTCH3 protein. All carriers were heterozygotes, giving a population frequency of 1 in 452, about 100-fold higher than expected based on CADASIL prevalence estimations of 2-5 in 100,000.[7]

### Distribution of *NOTCH3* pathogenic variants

The distribution of variants is shown in **Figure 1** and **Supplementary Table 2**. In total, 67 unique *NOTCH3* pathogenic variants were identified, of which 36 were observed in only one individual. The identified variants were spread across 22 different exons between exons 2 and 24 of the *NOTCH3* gene (except exon 16; **Figure 1**). They affected 31 EGFR domains, involving all EGFR domains except 7, 21 and 34. Variants were predominantly located towards the distal end of NOTCH3 protein; only eleven participants had a *NOTCH3* variant located in EGFR domains 1-6 (**Figure 1**). More than half of the variants (39 of 67, 58.2%) involved the swapping of cysteine and arginine residues.

**Figure 1.**
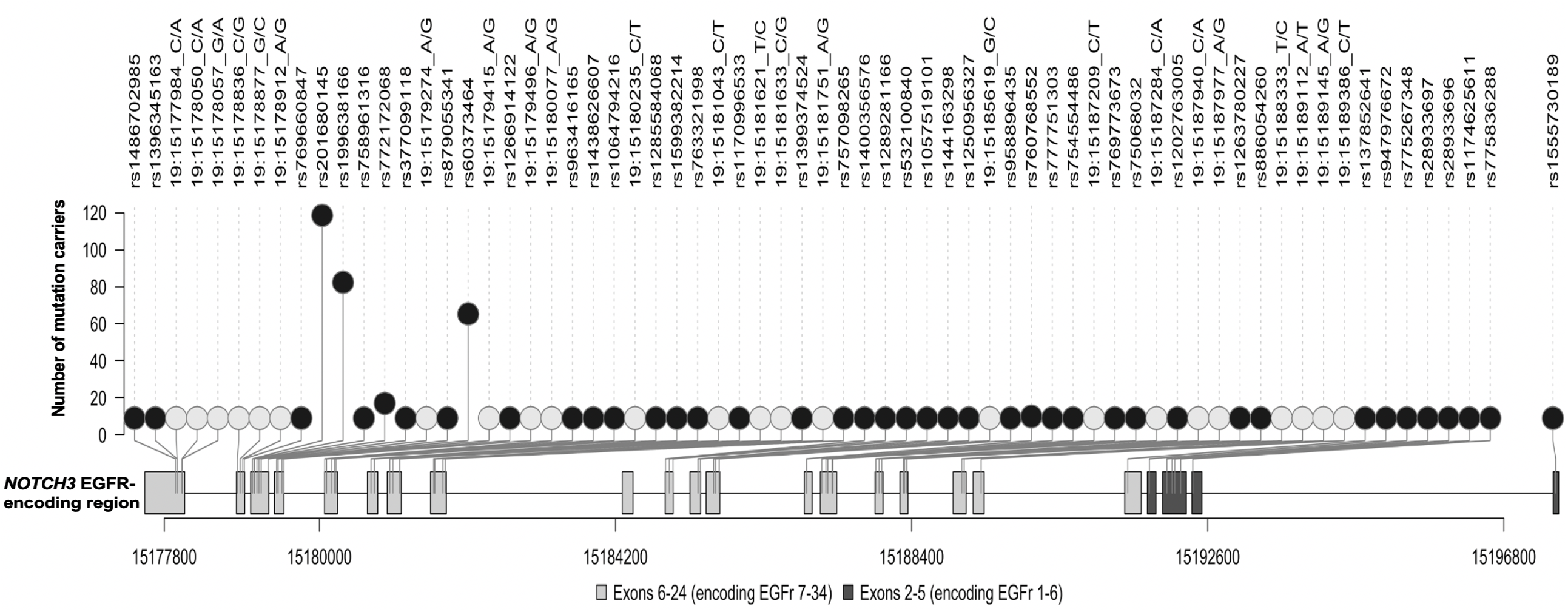
Lolliplot showing the distribution of distinct pathogenic variants in UK Biobank across the reverse strand of the NOTCH3 gene. Lighter circles represent variants without rsID; Darker circles represent variants with rsID; Lighter rectangular boxes represent exons that encode EGFRs 7-34; Darker rectangular boxes represent exons that encode EGFRs 1-6.

Sixty-five different variants were found in 378 White participants, three in forty-one individuals of Asian or Asian British ancestry, three in seven Black or Black British participants, one variant in a Chinese participant, one in four mixed ethnicity individuals, and four in other ethnic groups. Ten of the unique variants occurred in non-White subjects, and three were not observed in White individuals (p.Cys579Tyr, p.Gly861Cys and p.Cys1250Arg).

### *NOTCH3* variants were not associated with vascular risk factors, blood biochemistry, or cognition

The demographic and risk factor profile of cases with and without variants are shown in **Table 1**. There were no significant differences between groups, except that the variant group had more males and less White subjects. *NOTCH3* variants were not associated with vascular risk factors, blood biochemistry markers, the Framingham cardiovascular risk score or cognitive function (**Supplementary Figure 2**).

**Table 1.**
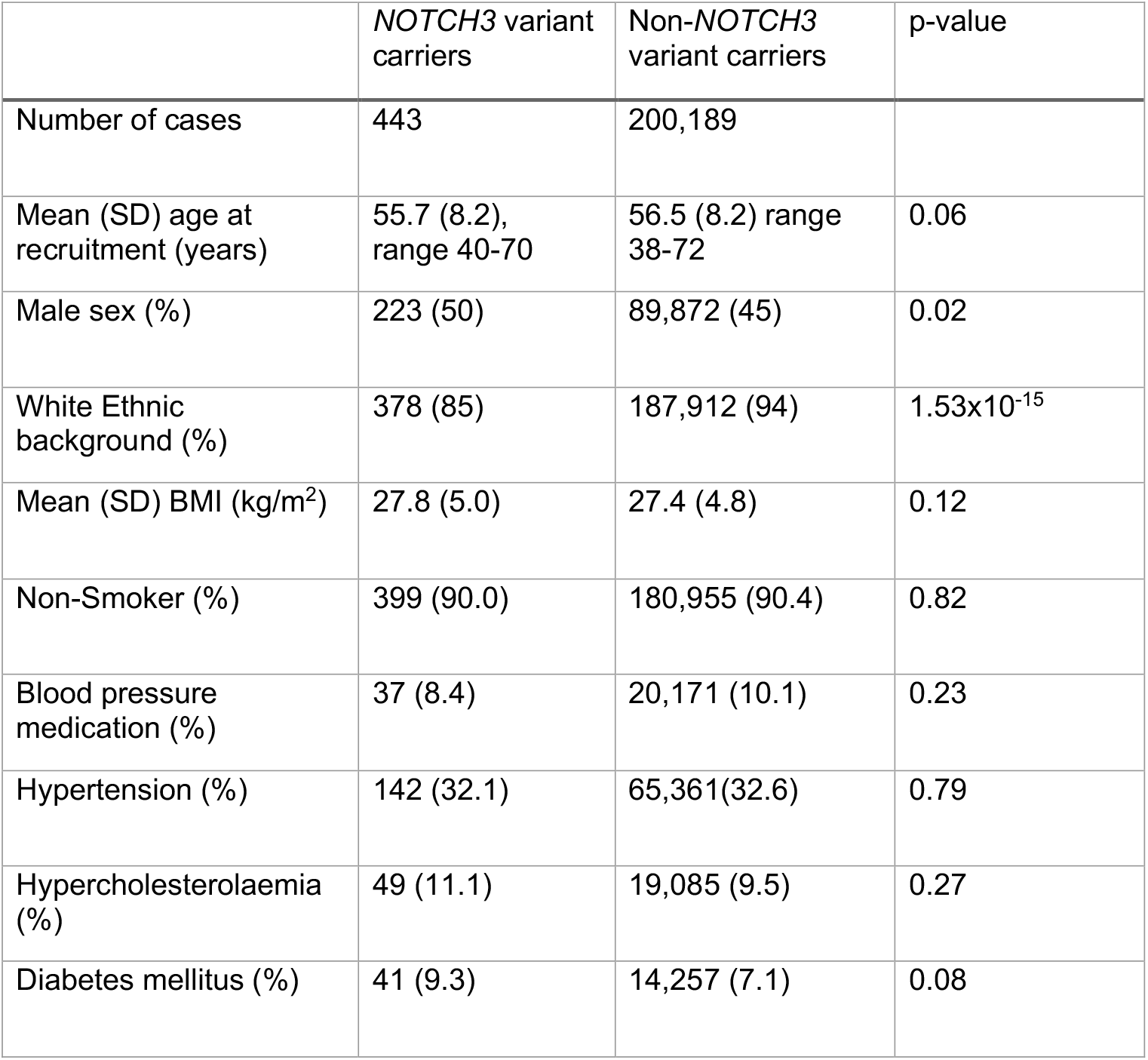
Comparison of the demographic and risk factor profile for cases with and without *NOTCH3* variants. The p-values for categorical and continuous variables were calculated by the Pearson’s Chi-squared test and two-sample t-test, respectively. BMI. Body mass index; SD, standard deviation.

### *NOTCH3* variant carriers were at an increased risk of stroke and vascular dementia

Presence of *NOTCH3* variants was associated with a two-fold increase in the odds of stroke (OR: 2.33, 95% CI: 1.50-3.45, p=0.0003). Vascular dementia (OR: 5.03, 1.67-11.50, p=0.007) was also more frequent in variant carriers. All dementia cases in *NOTCH3* carriers were of vascular origin (and occurred after enrolment into UK Biobank). All-cause dementia was not significantly associated with the presence of *NOTCH3* variants (OR: 2.13, 0.71-4.82, p=0.16). There was a borderline significant increase in epilepsy (OR:1.92, 1.01-3.29, p=0.048). Presence of a variant was associated with an increased family history of stroke (OR:1.41, 1.06-1.85, p=0.02). No significant associations were found with migraine (OR: 1.41, 0.94-2.03, p=0.09) or depression (OR: 0.88, 0.56-1.32, p=0.55) (**Figure 2**).

**Figure 2.**
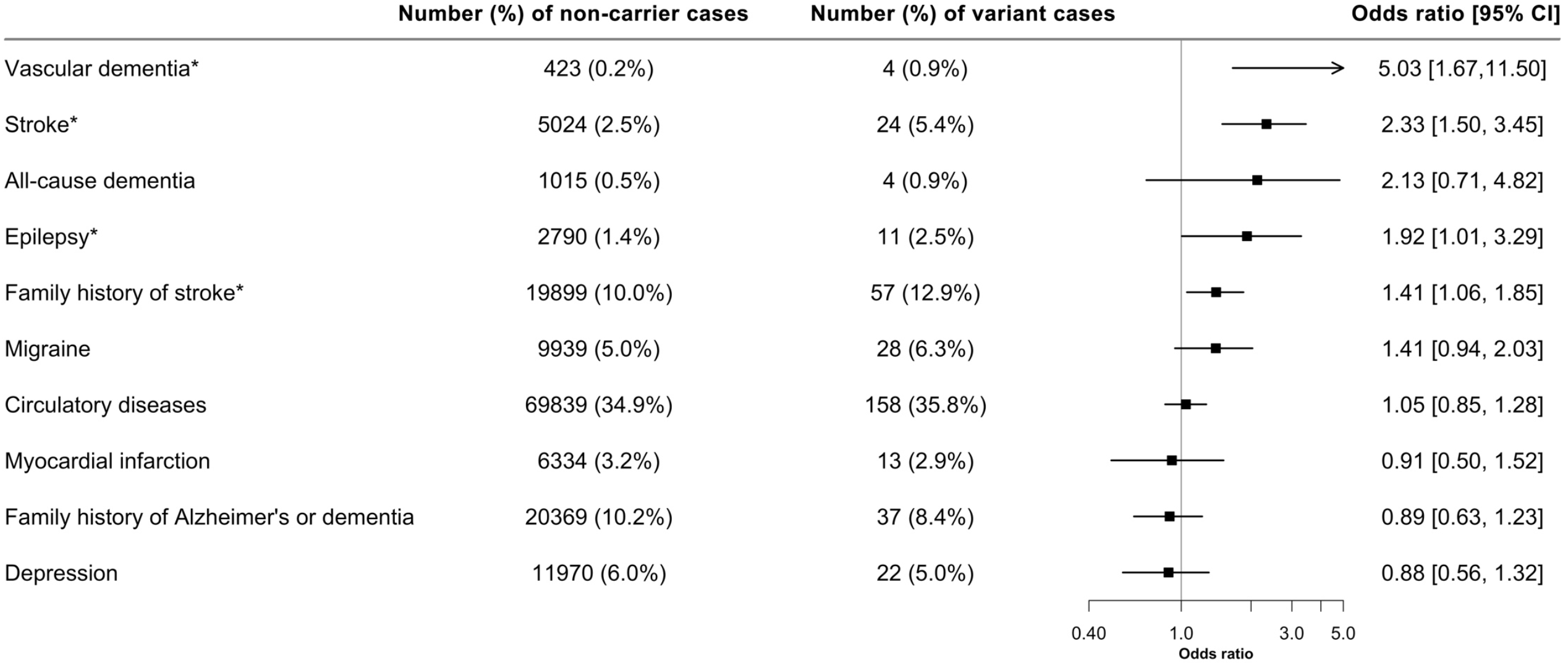
Forest plot showing the association of *NOTCH3* variant with different diagnoses. Family history and prevalence of disease cases were analysed with the presence of *NOTCH3* variant through logistic regression. Firth’s correction was applied to all the regression models. N = 200,632. CI, confidence interval; *, significance at p < 0.05.

The risk of both stroke and vascular dementia was higher with variants in EGFRs 1-6 compared to EGFRs 7-34 (**Figure 3**). The risk of stroke was thirteen-fold higher (OR 13.57, 2.46-53.16, p=0.006) and over double (OR 2.18, 1.37-3.26, p=0.001) that of variant-free individuals, respectively for variants in EGFRs 1-6 and 7-34, with findings for vascular dementia risk mirroring this.

**Figure 3.**
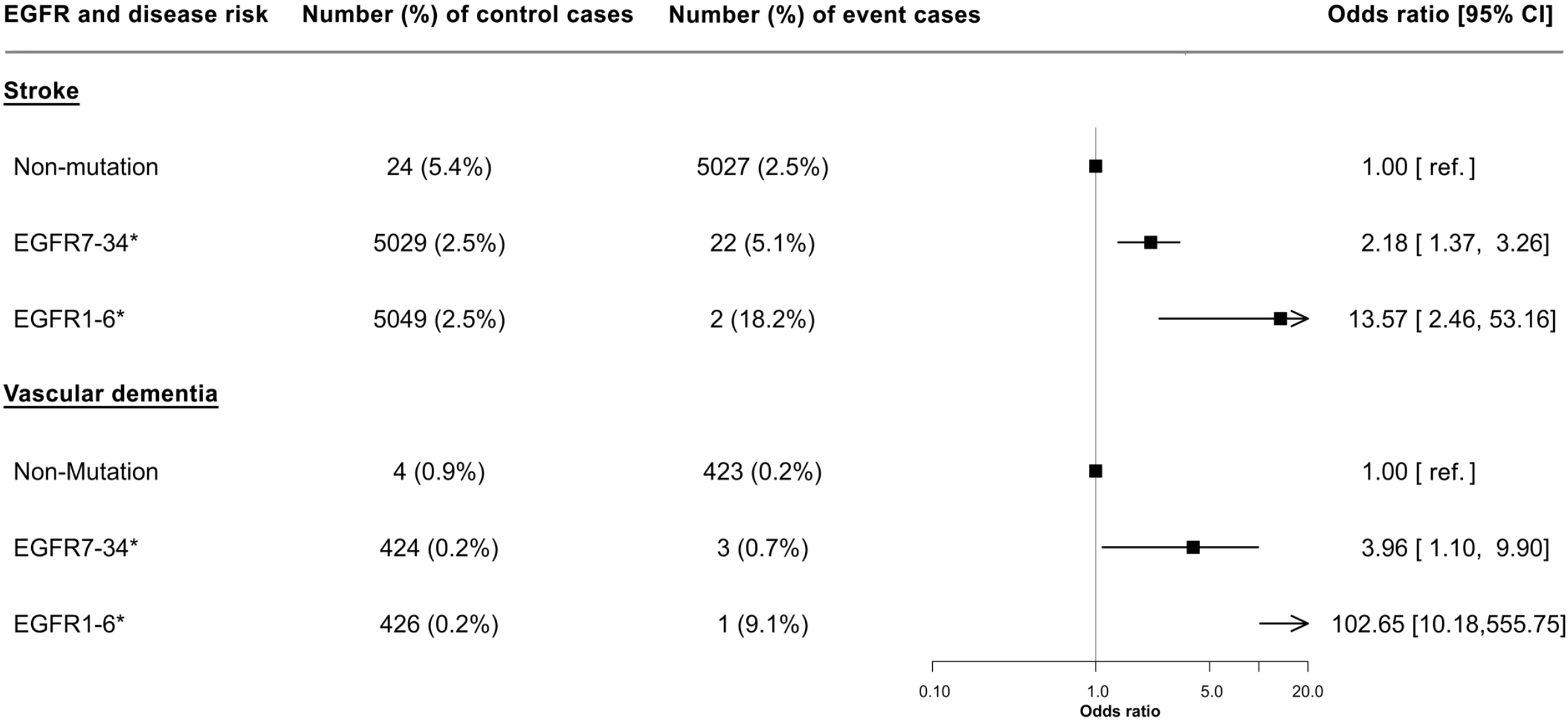
Forest plot showing the effect of *NOTCH3* variant location on the odds of stroke or vascular dementia. The location of variant was stratified by EGFRs 7-34 and EGFRs 1-6; their effect on disease risk was relative to that without the variant. Firth’s correction was applied to all the regression models. N = 200,632. CI, confidence interval; *, significance at p < 0.05.

Similar associations were observed when the same analyses were performed for incident cases of outcomes i.e., those that had occurred from the point of recruitment into UK Biobank to the latest follow-up in March 2019 (**Supplementary Figures 3 and 4**).

### *NOTCH3* variant carriers showed signs of brain MRI abnormalities

*NOTCH3* variants were associated with an increase in WMH volume (standardised difference 0.52, 95% CI: 0.29-0.75, p=1.0×10^−5^) and DTI-PSMD (standardised difference: 0.71, 0.39-1.03, p=1.0×10^−5^) but not with brain volume.

MRI scans from 47 *NOTCH3* variant carriers and 148 matched controls were visually rated and compared (**Table 2, Figure 4 and Supplementary Figure 5**). Image sequences available were T1-weighted (47 cases, 148 controls), T2 FLAIR (45 cases, 148 controls) and SWI (44 cases, 148 controls). *NOTCH3* variants were associated with an increased risk of the presence of lacunes (OR: 4.83, 2.00-12.06, p=0.0007) and cerebral microbleeds (OR: 3.61,1.69-7.76, p=0.0009). There were also significant associations with the numbers of lacunes (standardised difference: 0.56, 0.23-0.89, p=0.001) and of cerebral microbleeds (standardised difference: 0.39, 0.05-0.74, p=0.03).

**Figure 4.**
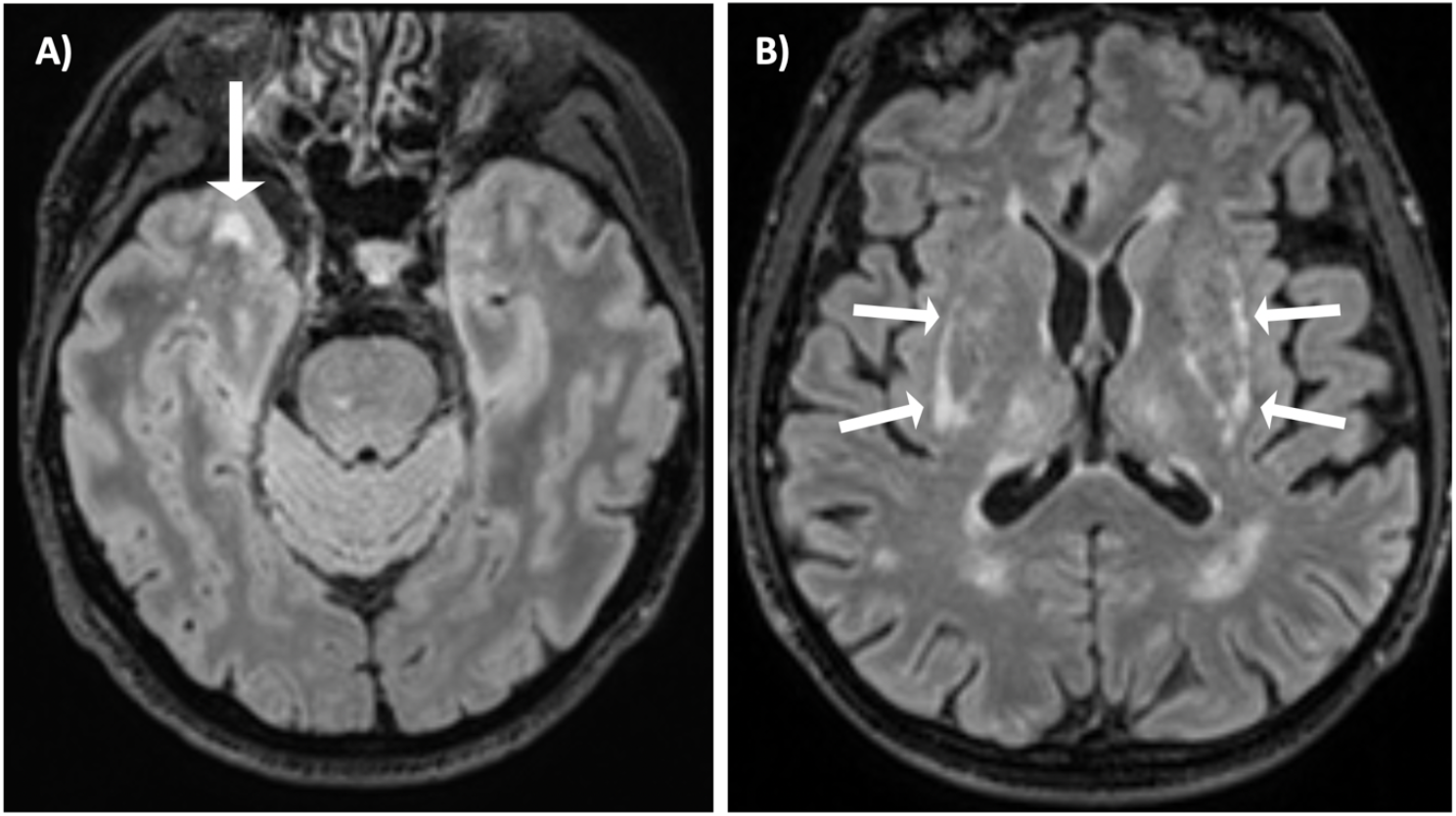
Axial T2 FLAIR brain MRI of 2 subjects with a pathogenic *NOTCH3* variant in UK Biobank. (A) A woman in her 50s, carrying a p.Arg1231Cys variant that affects EGFR domain 31, with WMH involving the right anterior temporal lobe (arrow). (B) A man in his 60s, carrying a p.Arg578Cys variant that affects EGFR domain 14, with WMH involving the external capsule bilaterally (arrows).

**Table 2.** Comparison of WMH distribution on the modified Schelten’s scale between *NOTCH3* variant carriers and non-carriers. Table showing the effect of the presence of *NOTCH3* variants on the odds of developing any WMH in each brain region.

| Brain regions | Number (%) of non-carrier cases | Number (%) of mutation cases | Odds ratio [95% CI] | P-value |
| --- | --- | --- | --- | --- |
| Periventricular white matter | 108 (73%) | 36 (80%) | 1.46 [0.64,3.62] | 0.38 |
| Frontal white matter | 119 (80%) | 40 (89%) | 1.50 [0.55,4.74] | 0.44 |
| Parietal white matter | 79 (53%) | 34 (76%) | 2.79 [1.29,6.45] | 0.01 |
| Occipital white matter | 45 (30%) | 20 (44%) | 1.77 [0.86,3.61] | 0.12 |
| Anterior temporal white matter | 12 (9%) | 17 (38%) | 6.92 [2.91,17.29] | <0.001 |
| Posterior temporal white matter | 45 (30%) | 14 (31%) | 1.03 [0.48,2.19] | 0.93 |
| Corpus callosum | 7 (5%) | 5 (11%) | 2.41 [0.69,7.98] | 0.16 |
| Caudate | 1 (1%) | 3 (7%) | 7.05 [1.15,73.15] | 0.04 |
| Putamen | 16 (11%) | 9 (20%) | 2.00 [0.80,4.88] | 0.14 |
| Globus pallidus | 4 (3%) | 0 (0%) | 0.33 [0.00,3.17] | 0.40 |
| Thalamus | 4 (3%) | 6 (13%) | 4.38 [1.25,16.21] | 0.02 |
| Internal capsule | 3 (2%) | 2 (4%) | 1.79 [0.30,8.71] | 0.49 |
| External capsule | 9 (6%) | 17 (38%) | 12.44 [4.67,37.37] | <0.001 |
| Cerebellum | 4 (3%) | 3 (7%) | 1.99 [0.41,8.15] | 0.37 |
| Mesencephalon | 3 (2%) | 2 (4%) | 3.30 [0.50,24.23] | 0.20 |
| Pons | 25 (17%) | 11 (24%) | 1.65 [0.71,3.70] | 0.24 |
| Medulla | 2 (1%) | 0 (0%) | 0.69 [0.01,7.02] | 0.78 |

### *NOTCH3* variants were associated with a CADASIL-like distribution of WMH

The presence of *NOTCH3* variants was correlated with the spatial distribution and extent of WMH using the modified Schelten’s scale. First, we examined the distribution of the presence of any WMH in each brain region (**Table 2**). *NOTCH3* variants were most strongly associated with any WMH in two areas previously reported to be predilection sites for WMH in CADASIL,[25] the anterior temporal lobe (OR 6.92, 2.91,17.29 p <0.001) and external capsule (OR 12.44, CI 4.67-37.37, p <0.001). Typical example scans are shown in **Figure 4**. Secondly, we examined the severity of WMH in each region, as assessed on the Schelten’s scale. *NOTCH3* variants were associated with higher aggregate WMH score (standardised difference: 0.54, 0.24-0.84, p=5×10^−4^) and higher individual scores for eight of the seventeen brain regions examined, including the periventricular region, frontal lobe, parietal lobe, occipital lobe, anterior temporal lobe, caudate, thalamus and external capsule (**Figure 5**). Concordantly, the differences between variant carriers and non-carriers were most marked for the anterior temporal lobe and the external capsule. All these associations remained significant after accounting for multiple comparisons using a false-discovery rate of 5% with the Benjamini-Hochberg procedure.

**Figure 5.**
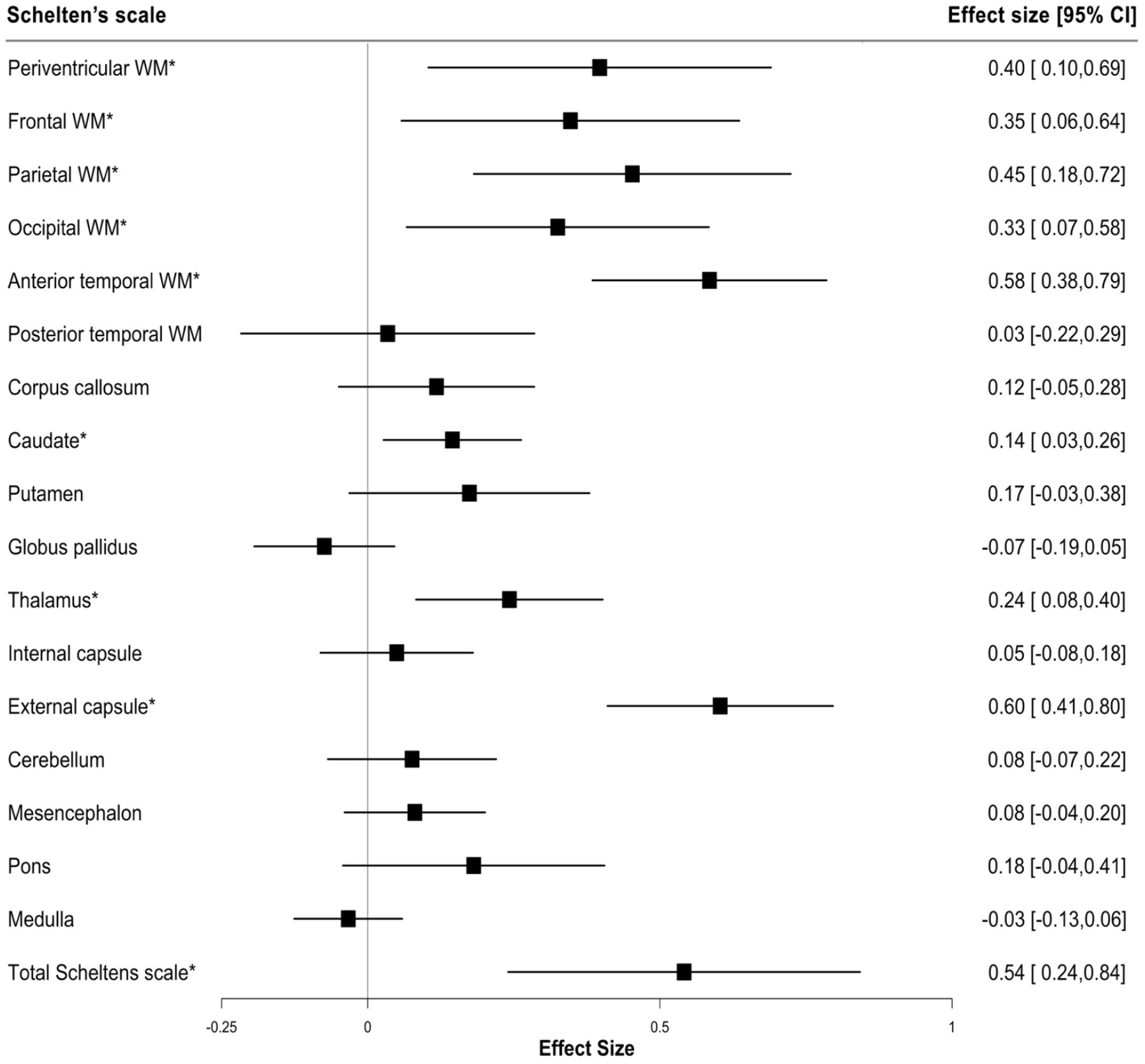
Forest plot showing the severity of WMH in different brain regions. Inversely transformed measures of the Schelten’s scale was analysed with the presence of *NOTCH3* variant through linear regression. A total of 193 participants were included in the regression analyses, of which 45 were variant carriers and 148 were non-carriers. DWH, deep white matter hyperintensities; PVH, periventricular hyperintensities; WM, white matter; CI, confidence interval; *, significance p < 0.05.

### *NOTCH3* associations with ischaemic stroke were independent of polygenic risk

*NOTCH3* carriers had a three-fold increased risk of ischaemic stroke (OR 3.08, 1.81-5.23, p=0.01). while a 1 SD increase in ischaemic stroke PRS increased the odds by 26% (OR:1.26, 1.21-1.32, p<0.01). We calculated presence of a *NOTCH3* mutation therefore conferred the same risk as a 4.8SD increase in PRS (**Figure 6**). Conscious of substantial uncertainty in our estimated association for *NOTCH3* status and ischaemic stroke we calculated more conservative estimates using the lower bound of 95%CI for *NOTCH3* with the OR (conservative *NOTCH3*) as well as upper bound of the 95%CI for the association with the PRS (most conservative). These placed carriers of *NOTCH3* variants as having a polygenic risk equivalent to the upper 1.6% of the population. We observed no interaction between *NOTCH3* carrier status and PRS (**Supplementary Figure 6**).

**Figure 6.**
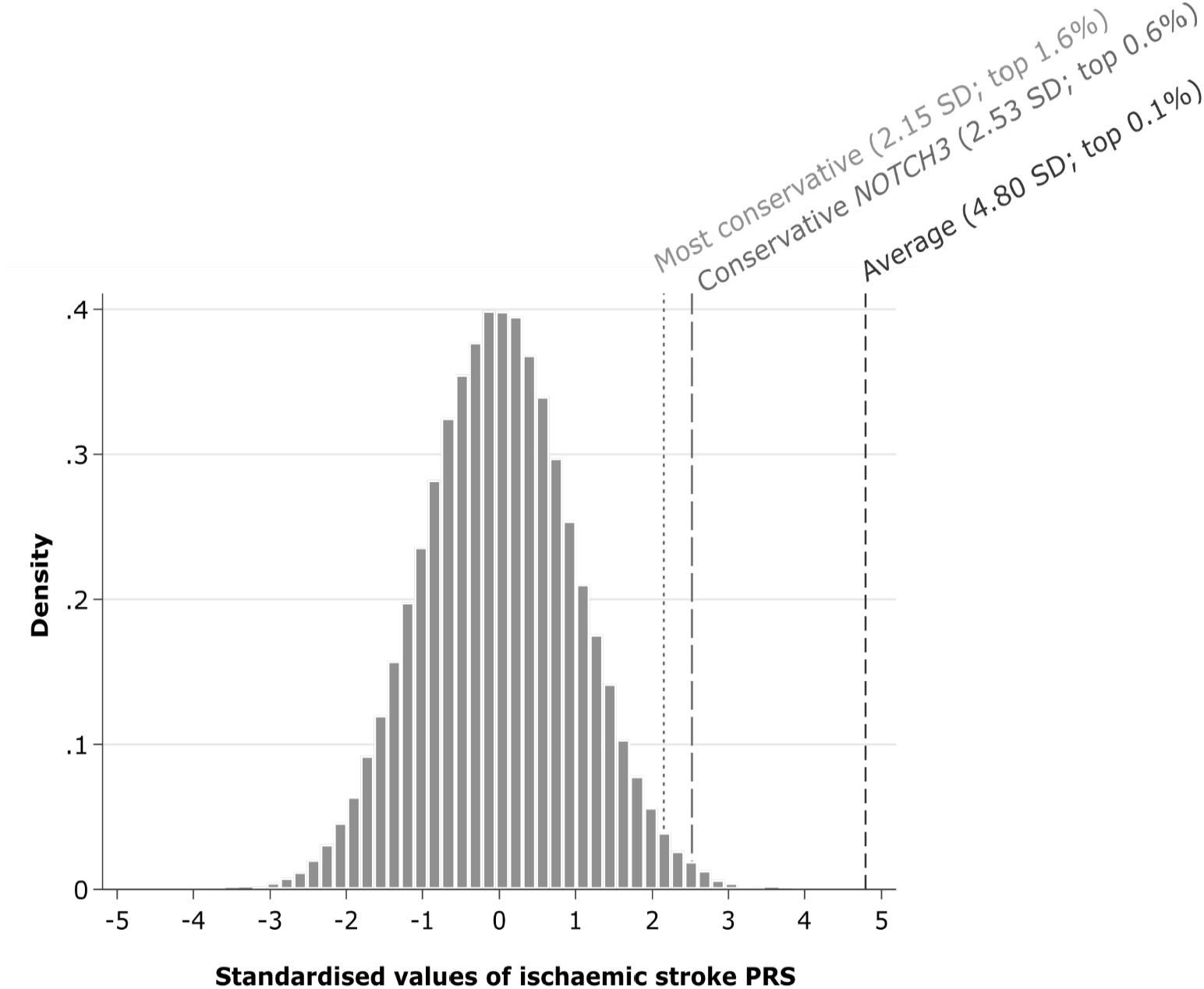
Estimated polygenic risk equivalent for the association of NOTCH3 status with ischaemic stroke. Average = ratio of the log-odds ratio of models of *NOTCH3* status and a 1 standard deviation increase in polygenic risk score for ischaemic stroke; conservative *NOTCH3* = ratio of the lower 95% confidence interval of the log-odds ratio for *NOTCH3* status and the log-odds ratio for a 1 standard deviation increase in polygenic risk score for ischaemic stroke; most conservative = ratio of the lower 95% confidence interval of the log-odds ratio for *NOTCH3* status and the upper 95% confidence interval of the log-odds ratio for a 1 standard deviation increase in polygenic risk score for ischaemic stroke. Population percentages calculated using a 1-sided Z-score.

## DISCUSSION

In this study of 200,632 individuals, we show that *NOTCH3* variants are much more common than expected in the general population, being present in 1 in 450 individuals. We demonstrated that these ‘asymptomatic’ variants are associated with an increased risk of both stroke and vascular dementia, and with MRI features of SVD, including lacunar infarcts, WMH, diffuse white matter damage on DTI, and cerebral microbleeds. Taken together, these results suggest that typical CADASIL variants are common in the general population, and account for a significant burden of apparently “sporadic” stroke and dementia.

CADASIL is reported to be rare, with an estimated disease prevalence of 4 per 100,000 individuals in UK populations.[5,6] In contrast, recent studies have demonstrated an increased frequency of cysteine-changing *NOTCH3* variants in population databases,[7,9] but these have been anonymised databases making it impossible to determine whether these variants associate with clinical disease. A very recent report from the United States Geisinger database suggested such variants associated with an increased risk of stroke and MRI features of SVD, although no association was found with dementia.[8] Our findings provide robust evidence from a large population-based study of over 200,000 individuals that common *NOTCH3* variants are associated with symptomatic cerebrovascular disease in the general population. We demonstrated that they conferred a risk equivalent to that in the top 1.6% of individuals as determined by a polygenic risk score, with the risks from common polygenic variant and *NOTCH3* variants being independent.

In addition to an association with stroke we demonstrate a strong association with vascular dementia. All cases of dementia occurring in patients with *NOTCH3* variants were of vascular; no association was found with all-cause dementia, reflecting the fact that many cases of dementia in older individuals are due to non-vascular pathologies. We also found trends towards other clinical features of CADASIL being more common with these variants with a marginally significant association with epilepsy, and a non-significant trend towards increased risk of migraine.

In addition to looking at overall brain MRI measures, we analysed the distribution of white matter lesions in individuals with *NOTCH3* variants, compared with matched controls. The greatest increase in WMH was in two areas known to be particularly affected in CADASIL, namely the anterior temporal lobes and external capsule.[25] This suggests that the *NOTCH3* variants are not merely associated with stroke, but also with a specific CADASIL-like phenotype in the general population. Although the phenotype of the variant carriers is much milder in the UK Biobank cohort, the distribution of white matter changes is similar to that seen in CADASIL patients.

It remains uncertain why some patients with *NOTCH3* variants develop the CADASIL phenotype, while others appear asymptomatic. One emerging factor is variant position. Within CADASIL cohorts, variants in EGFRs 1-6 have been associated with earlier stroke onset than variants in EGFRs 7-34.[10] In previous population studies, variants in EGFRs 7-34 were much more common,[10] and we confirmed this in UK Biobank. Furthermore, we found that although variants in EGFRs 1-6 were associated with higher risk of stroke and vascular dementia in the general population, more distal variants in EGFRs 7-34 were still associated with risk of both conditions. Nonetheless, variant site fails to account for much of the phenotypic heterogeneity. Vascular risk factors, particularly smoking and hypertension, have been shown to be associated with an earlier onset of stroke within CADASIL families but only account for a small amount of variability.[26,27] Family studies have suggested that as much of 60% of the heritability of WMH lesion volume is accounted for by yet undetermined modifier genetic factors outside the *NOTCH3* gene, but the nature of these remains undetermined.[28]

Our study has a number of strengths. Firstly, it included data from over 200,000 individuals with exome sequencing. Secondly, we correlated the genotyping with extensive clinical information, including prospectively collected information obtained from linked electronic health records. This allowed us to identify associations not only with disease prevalence but also incident disease occurring during prospective follow-up. This revealed very similar associations with stroke and vascular dementia to that observed in the prevalence data. Thirdly, all scans had been performed on one of the two identical MR scanners using identical sequences improving consistency and quality.

It also has limitations. Diseases like dementia can sometimes be misclassified or underrepresented by the ICD codes in health records. Our analysis was in predominantly White populations. CADASIL has been shown to be present in all ethnic groups in which it is being looked for, and we have no reason to suspect the associations reported are different in other ethnic groups. However, frequency of *NOTCH3* variants may differ between ethnic groups, and initial data suggests that it is higher in Far Eastern populations including Taiwan.[29] Pertinently, a recent report suggested typical CADASIL variants may predispose to multifactorial late onset stroke in Taiwan.[30]

In conclusion, our study shows that typical cysteine changing *NOTCH3* variants are common in the general population and these variants are associated with increased risk of both stroke and vascular dementia, and with MRI markers of SVD including WMH and lacunar infarcts. This demonstrates that genetic variation in the *NOTCH3* gene accounts for a much greater proportion of stroke in the general population than previously thought. Although the magnitude of risk for a *NOTCH3* variant on stroke and dementia partly depends on variant site, modifiers outside the *NOTCH3* gene appear to be important. Further research is required to identify these. Finally, our study emphasises that the boundaries between monogenic and polygenic stroke are not as distinct as previously believed.

## Data Availability

Data from UK Biobank are available to bona fide researchers on application. This analysis was performed under UK Biobank application number 36509.

## CONTRIBUTORS

B.C., S.B. and H.S.M. conceived and designed the study. B.C., E.H., S.G. and S.B. extracted and analysed the genetic and phenotypic data. B.C., S.N. and D.T. downloaded and analysed the neuroimaging data. B.C., S.B. and H.S.M. interpreted the data. B.C. and H.S.M. wrote the first draft of the manuscript. All authors contributed to writing the final version of the manuscript.

