## Supplementary for "*NOTCH3* variants are common in the general population and associated with stroke and vascular dementia: an analysis of 200,000 participants"

**SUPPLEMENTAL MATERIAL**
**(2 tables and 6 figures)**

### Supplementary table 1. The code list used for the retrieval of disease records.

| Biobank Code Text | Code type | Code | Date of last linkage |
|---|---|---|---|
| Stroke | UK Biobank algorithmically defined stroke | 42007 | March 2019 |
| | UK Biobank self-report | Field 20002 Codes 1081,1086,1491,1583 | May 2020 |
| | ICD9 for hospital and death record retrieval | 430, 431, 434, 436 | May 2020 |
| | ICD10 for hospital and death record retrieval | I60, I61, I63, I64 | May 2020 |
| Family history of stroke | Illness of father | Field 20107 Code 2 | May 2020 |
| | Illness of mother | Field 20110 Code 2 | May 2020 |
| All-cause dementia | UK Biobank algorithmically defined dementia | 42019 | March 2019 |
| Vascular dementia | UK Biobank algorithmically defined vascular dementia | 42023 | March 2019 |
| | ICD9 for hospital and death record retrieval | 290.2,2903,2904,2912,2941,3310,3311,3312, 3315 | May 2020 |
| | ICD10 for hospital and death record retrieval | F00, F01, F02, F03, G30, A81.0, F05.1,F10.6,G31.0,G31.1,G31.8,I67.3 | May 2020 |
| Family history of Alzhiemer's or dementia | Illness of father | Field 20107 Code 10 | May 2020 |
| | Illness of mother | Field 20110 Code 10 | May 2020 |
| Migraine | UK Biobank algorithmically defined migraine | 131053 | March 2019 |
| | UK Biobank self-report | Field 20002 Code 1265 | May 2020 |
| | ICD9 for hospital record retrieval | 346 | May 2020 |
| | ICD10 for hospital record retrieval | G43 | May 2020 |
| Epilepsy | UK Biobank algorithmically defined epilepsy | 131049 | March 2019 |
| | UK Biobank self-report | Field 20002 Code 1264 | May 2020 |
| | ICD9 for hospital and death record retrieval | 345 | May 2020 |
| | ICD10 for hospital and death record retrieval | G40 | May 2020 |
| Depression | Touchscreen Q&A | Fields 20124 and 20125 | May 2020 |
| | ICD9 for hospital record retrieval | 311 | May 2020 |
| | ICD10 for hospital record retrieval | F32,F33 | May 2020 |
| Myocardial infarction | UK Biobank algorithmically defined myocardial infarction | 42001 | March 2019 |
| | UK Biobank self-report | Field 20002 Code 1075 | May 2020 |
| | ICD9 for hospital and death record retrieval | 410, 411, 412, 436 | May 2020 |
| | ICD10 for hospital and death record retrieval | I21, I22, I23, I24.1,I25.2 | May 2020 |
| Circulatory diseases | ICD9 for hospital record retrieval | 39, 40, 41, 42, 43,44, 45 | May 2020 |
| | ICD10 for hospital record retrieval | I0, I1, I2, I3, I4, I5, I6, I7, I8, I9 | May 2020 |

**Supplementary table 2. Sixty-seven distinct cysteine altering *NOTCH3* variants identified in UK Biobank.** All the variants lead to an uneven number of cysteine residues in the EGFR domains of the NOTCH3 protein.

| Protein change | Genomic change | Codon change | Exon | EGFR domain | Frequency in UK Biobank | Ethnicities of variant carriers in UK Biobank |
|---|---|---|---|---|---|---|
| p.Arg54Cys | g.15197537G>A | c.160C>T | 2 | 1 | 1 | White (1) |
| p.Arg110Cys | g.15192389G>A | c.328C>T | 3 | 2 | 1 | White (1) |
| p.Arg141Cys | g.15192218G>A | c.421C>T | 4 | 3 | 3 | White (3) |
| p.Arg169Cys | g.15192134G>A | c.505C>T | 4 | 4 | 1 | White (1) |
| p.Arg182Cys | g.15192095G>A | c.544C>T | 4 | 4 | 3 | White (3) |
| p.Arg207Cys | g.15192020G>A | c.619C>T | 4 | 5 | 1 | White (1) |
| p.Tyr258Cys | g.15191774T>C | c.773A>G | 5 | 6 | 1 | Others (1) |
| p.Arg332Cys | g.15191466G>A | c.994C>T | 6 | 8 | 1 | White (1) |
| p.Cys360Tyr | g.15189386C>T | c.1079G>A | 7 | 9 | 1 | White (1) |
| p.Cys408Arg | g.15189145A>G | c.1222T>C | 8 | 10 | 1 | White (1) |
| p.Cys419Ser | g.15189112A>T | c.1255T>A | 8 | 10 | 1 | White (1) |
| p.Tyr465Cys | g.15188333T>C | c.1394A>G | 9 | 11 | 1 | White (1) |
| p.Ser476Cys | g.15188301T>A | c.1426A>T | 9 | 12 | 2 | White (2) |
| p.Gly481Cys | g.15188286C>A | c.1441G>T | 9 | 12 | 1 | White (1) |
| p.Cys504Arg | g.15187977A>G | c.1510T>C | 10 | 12 | 1 | White (1) |
| p.Cys516Phe | g.15187940C>A | c.1547G>T | 10 | 13 | 6 | White (6) |
| p.Arg532Cys | g.15187893G>A | c.1594C>T | 10 | 13 | 2 | White (2) |
| p.Cys554Phe | g.15187284C>A | c.1661G>T | 11 | 14 | 1 | White (1) |
| p.Arg558Cys | g.15187273G>A | c.1672C>T | 11 | 14 | 1 | White (1) |
| p.Arg578Cys | g.15187213G>A | c.1732C>T | 11 | 14 | 7 | White (6), Asian or Asian British (1) |
| p.Cys579Tyr | g.15187209C>T | c.1736G>A | 11 | 14 | 1 | Black or Black British (1) |
| p.Arg587Cys | g.15187186G>A | c.1759C>T | 11 | 15 | 4 | White (3), Black or Black British (1) |
| p.Arg607Cys | g.15187126G>A | c.1819C>T | 11 | 15 | 4 | White (4) |
| p.Arg640Cys | g.15186911G>A | c.1918C>T | 12 | 16 | 13 | White (8), Asian or Asian British (1), Chinese (1), Others (3) |
| p.Cys654Tyr | g.15185670C>T | c.1961G>A | 13 | 16 | 1 | White (1) |
| p.Ser671Cys | g.15185619G>C | c.2012C>G | 13 | 17 | 2 | White (2) |
| p.Arg680Cys | g.15185593G>A | c.2038C>T | 13 | 17 | 1 | White (1) |
| p.Arg717Cys | g.15185404G>A | c.2149C>T | 14 | 18 | 11 | White (11) |
| p.Arg728Cys | g.15185371G>A | c.2182C>T | 14 | 18 | 5 | White (5) |
| p.Arg767Cys | g.15185017G>A | c.2299C>T | 15 | 19 | 5 | White (5) |
| p.Arg785Cys | g.15184963G>A | c.2353C>T | 15 | 20 | 7 | White (7) |

| | | | | | | |
|---|---|---|---|---|---|---|
| p.Trp802Cys | g.15184910C>A | c.2406G>T | 15 | 20 | 3 | White (3) |
| p.Gly861Cys | g.15181787C>A | c.2581G>T | 17 | 22 | 1 | Black or Black British (1) |
| p.Cys873Arg | g.15181751A>G | c.2617T>C | 17 | 22 | 1 | White (1) |
| p.Cys910Tyr | g.15181639C>T | c.2729G>A | 17 | 23 | 2 | White (2) |
| p.Cys912Ser | g.15181634A>T | c.2734T>A | 17 | 23 | 2 | White (2) |
| p.Tyr916Cys | g.15181621T>C | c.2747A>G | 17 | 23 | 3 | White (3) |
| p.Cys939Ser | g.15181140A>T | c.2815T>A | 18 | 24 | 2 | White (1) |
| p.Cys971Tyr | g.15181043C>T | c.2912G>A | 18 | 25 | 1 | White (1) |
| p.Cys986Arg | g.15180999A>G | c.2956T>C | 18 | 25 | 1 | White (1) |
| p.Cys1015Arg | g.15180780A>G | c.3043T>C | 19 | 26 | 1 | White (1) |
| p.Arg1031Cys | g.15180732G>A | c.3091C>T | 19 | 26 | 3 | White (3) |
| p.Cys1055Tyr | g.15180235C>T | c.3164G>A | 20 | 27 | 1 | White (1) |
| p.Cys1061Tyr | g.15180217C>T | c.3182G>A | 20 | 27 | 1 | White (1) |
| p.Arg1076Cys | g.15180173G>A | c.3226C>T | 20 | 27 | 1 | White (1) |
| p.Arg1100Cys | g.15180101G>A | c.3298C>T | 20 | 28 | 1 | White (1) |
| p.Cys1108Arg | g.15180077A>G | c.3322T>C | 20 | 28 | 1 | White (1) |
| p.Cys1110Arg | g.15179496A>G | c.3328T>C | 21 | 28 | 2 | White (2) |
| p.Cys1119Tyr | g.15179468C>T | c.3356G>A | 21 | 28 | 2 | White (2) |
| p.Cys1137Arg | g.15179415A>G | c.3409T>C | 21 | 29 | 1 | White (1) |
| p.Arg1143Cys | g.15179397G>A | c.3427C>T | 21 | 29 | 68 | White (66), Black or Black British (2) |
| p.Tyr1144Cys | g.15179153T>C | c.3431A>G | 21 | 29 | 2 | White (2) |
| p.Cys1157Arg | g.15179274A>G | c.3469T>C | 22 | 29 | 1 | White (1) |
| p.Arg1190Cys | g.15179175G>A | c.3568C>T | 22 | 30 | 9 | White (9) |
| p.Arg1201Cys | g.15179142G>A | c.3601C>T | 22 | 30 | 20 | White (19), Black or Black British (1) |
| p.Arg1210Cys | g.15179115G>A | c.3628C>T | 22 | 31 | 2 | White (1), Others (1) |
| p.Cys1222Gly | g.15179079A>C | c.3664T>G | 22 | 31 | 85 | White (85) |
| p.Arg1231Cys | g.15179052G>A | c.3691C>T | 22 | 31 | 121 | White (72), Mixed (4), Asian or Asian British (38), Black or Black British (1), Others (6) |
| p.Arg1242Cys | g.15178936G>A | c.3724C>T | 23 | 31 | 5 | White (5) |
| p.Cys1250Arg | g.15178912A>G | c.3748T>C | 23 | 32 | 1 | Asian or Asian British (1) |
| p.Cys1261Trp | g.15178877G>C | c.3783C>G | 23 | 32 | 1 | White (1) |
| p.Cys1275Ser | g.15178837A>T | c.3823T>A | 23 | 32 | 1 | White (1) |
| p.Arg1291Cys | g.15178057G>A | c.3871C>T | 24 | 33 | 1 | White (1) |
| p.Cys1293Phe | g.15178050C>A | c.3878G>T | 24 | 33 | 1 | White (1) |
| p.Cys1315Trp | g.15177983G>C | c.3945C>G | 24 | 33 | 1 | White (2) |
| p.Cys1315Phe | g.15177984C>A | c.3944G>T | 24 | 33 | 2 | White (1) |
| p.Cys1324Ser | g.15177958A>T | c.3970T>A | 24 | 33 | 1 | White (1) |

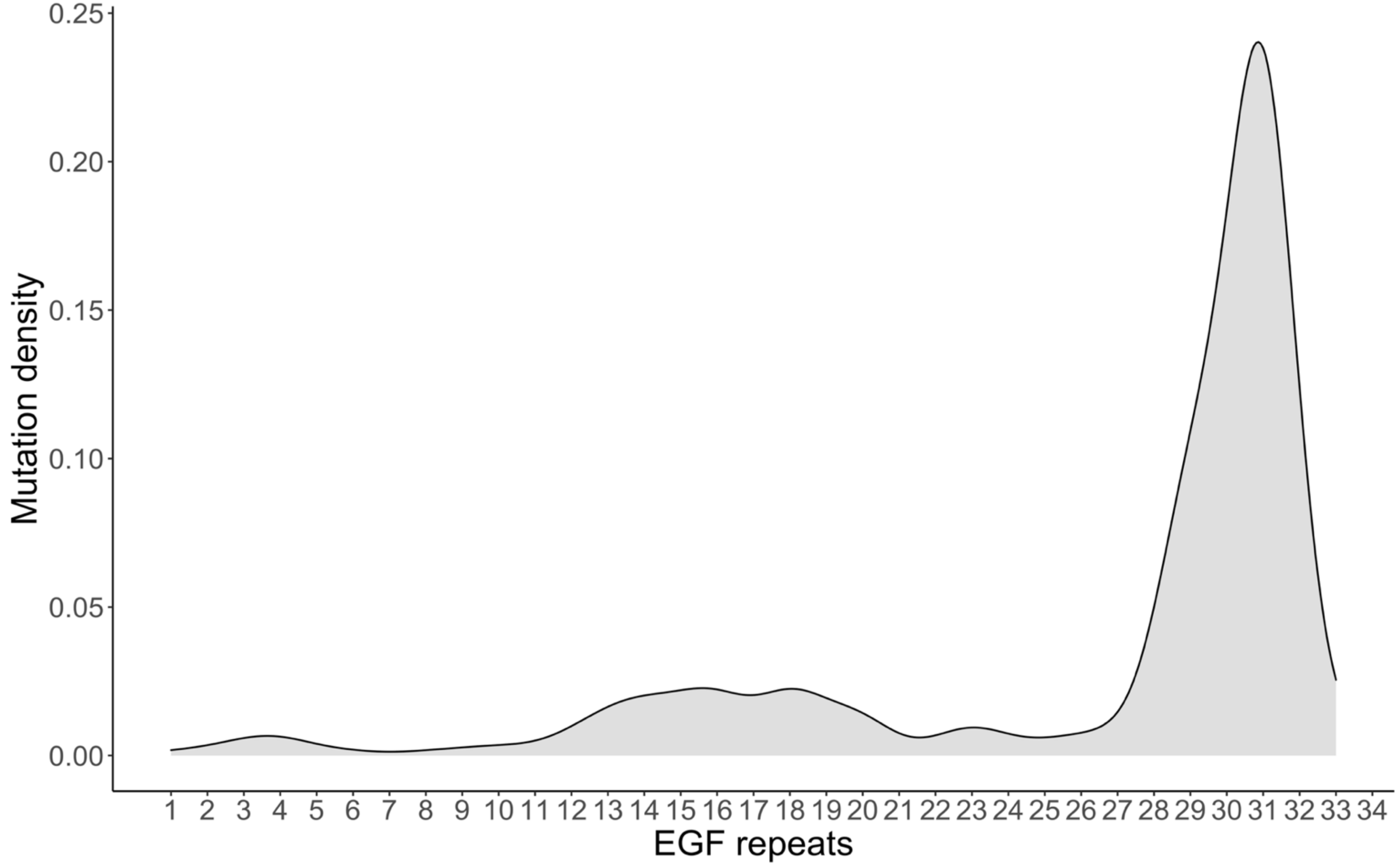


**Supplementary figure 1. Density plot showing the distribution of variants across the 34 EGFR domains of *NOTCH3***

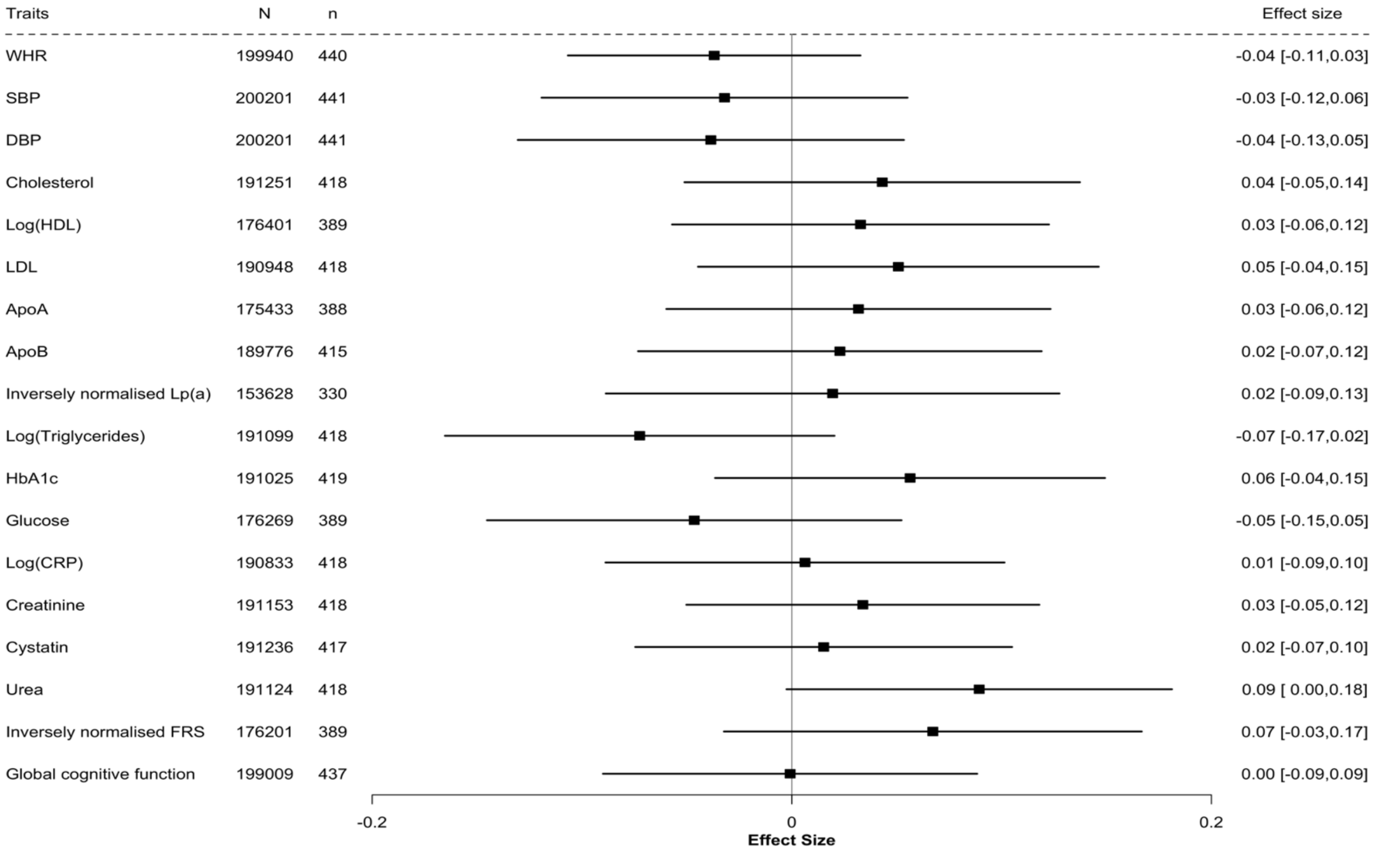


**Supplementary figure 2. Forest plot showing the standardised effect of *NOTCH3* variant on different clinical traits.** Measurements of blood pressure, blood biochemistry, Framingham risk score (FRS) and cognitive function were analysed with the presence of *NOTCH3* variant through linear regression. N, total number of participants included in regression analysis; n, number of variant carriers included in the analysis; CI, confidence interval; bp, blood pressure; HDL, high-density lipoprotein; LDL, low-density lipoprotein; ApoA, apolipoprotein a; ApoB, apolipoprotein b; Lpa, lipoprotein a; HbA1c, glycated haemoglobin.

| Disease incidence | Number (%) of non-carrier cases | Number (%) of variant cases | Odds ratio [95% CI] |
|---|---|---|---|
| Vascular dementia* | 189 (0.09%) | 4 (0.9%) | 11.13 [3.66,25.74] |
| Epilepsy* | 693 (0.3%) | 6 (1.4%) | 4.41 [1.81, 8.85] |
| Stroke* | 2001 (1.0%) | 12 (2.7%) | 2.98 [1.60, 5.04] |
| All-cause dementia | 925 (0.5%) | 4 (0.9%) | 2.34 [0.78, 5.32] |

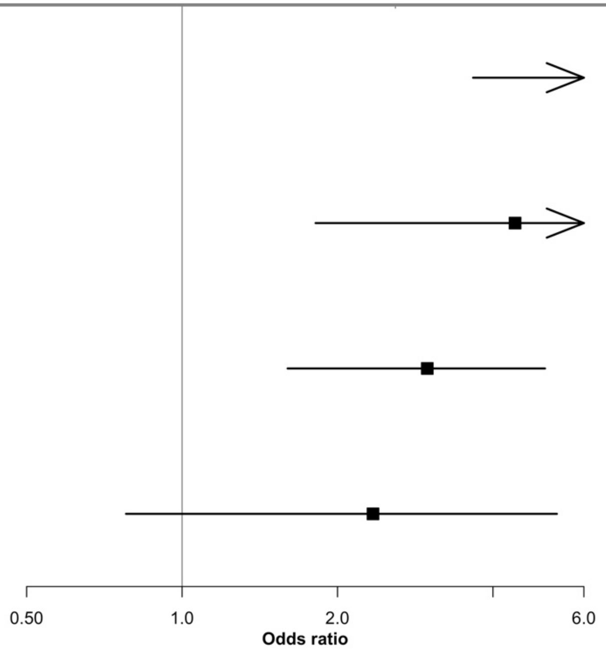


**Supplementary figure 3. Forest plot showing the effect of *NOTCH3* variants on the odds of incident cases of stroke, all-cause dementia, vascular dementia or epilepsy.** Firth's correction was applied to all the logistic regression models. Total number of participants included in the regression analyses (N) = 200,632. CI, confidence interval; *, significance at $p < 0.05$.

| EGFR and disease incidence | Number (%) of control cases | Number (%) of event cases | Odds ratio [95% CI] |
|---|---|---|---|
| **Stroke** | | | |
| Non-mutation | 12 (2.8%) | 2002 (1.0%) | 1.00 [ ref. ] |
| EGFR7-34* | 2002 (1.0%) | 12 (2.8%) | 3.05 [ 1.64, 5.16] |
| EGFR1-6 | 2014 (1.0%) | 0 (0.0%) | 5.28 [ 0.04, 43.83] |
| **Vascular dementia** | | | |
| Non-Mutation | 4 (0.9%) | 189 (0.09%) | 1.00 [ ref. ] |
| EGFR7-34* | 190 (0.09%) | 3 (0.7%) | 8.74 [ 2.42, 22.08] |
| EGFR1-6* | 192 (0.1%) | 1 (9.1%) | 248.42 [24.51,1369.11] |

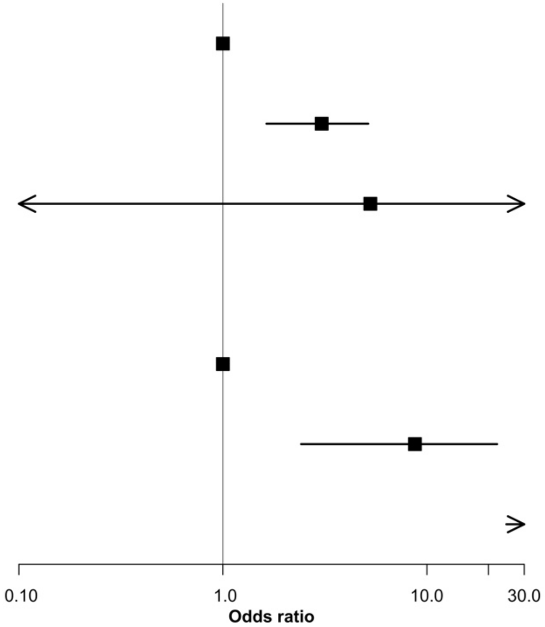


**Supplementary figure 4. Forest plot showing the effect of *NOTCH3* variant location on the odds of stroke or vascular dementia cases from the point of recruitment to last follow-up.** The location of variant was stratified by EGFR 7-34 and EGFR 1-6; their effect on disease risk was relative to that without the variant. Firth's correction was applied to all the regression models. Total number of participants included in the regression analyses (N) = 200,632. CI, confidence interval; *, significance at $p < 0.05$.

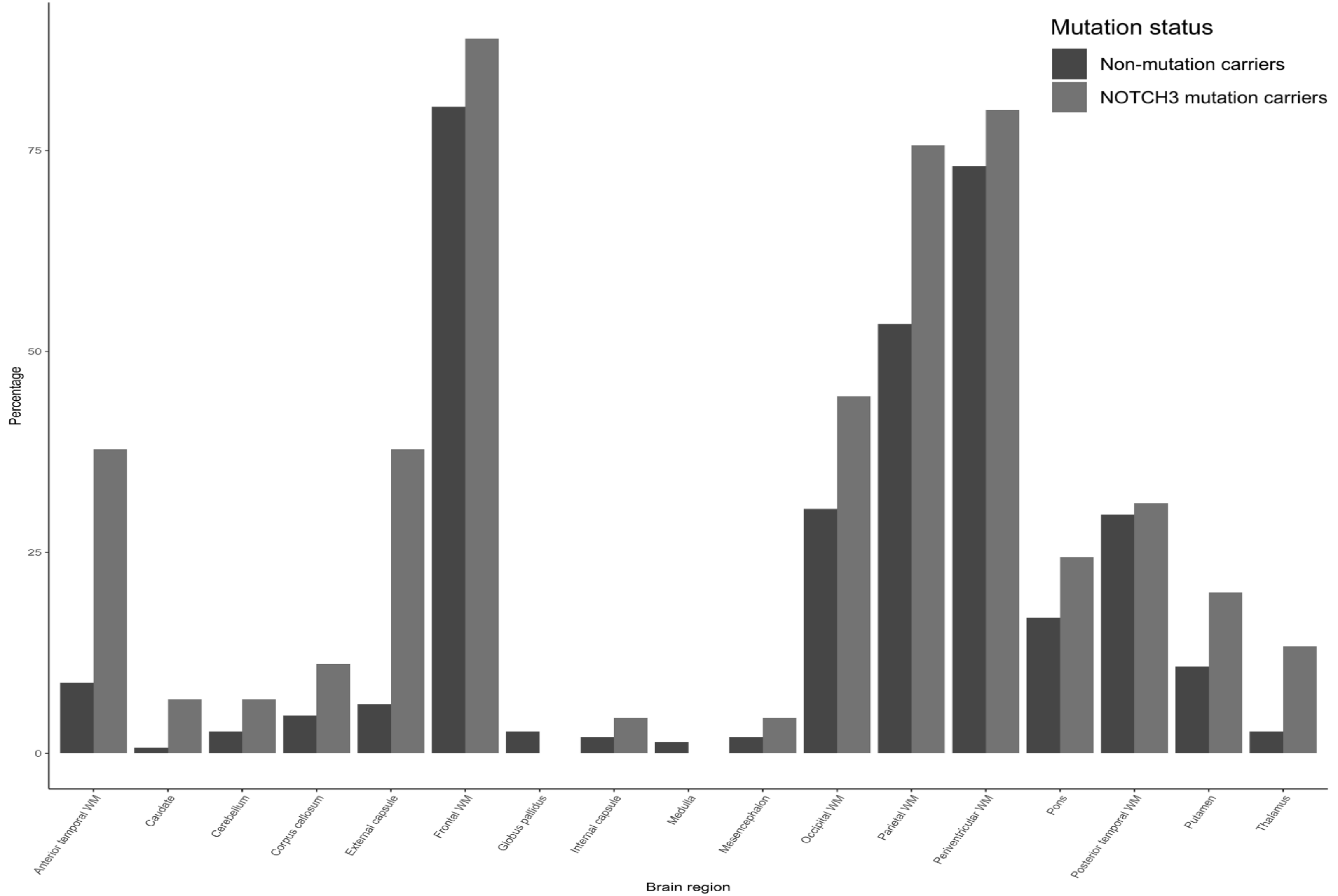


**Supplementary figure 5.** Bar chart showing the proportion of participants who had any WMH present in the each of the brain regions.

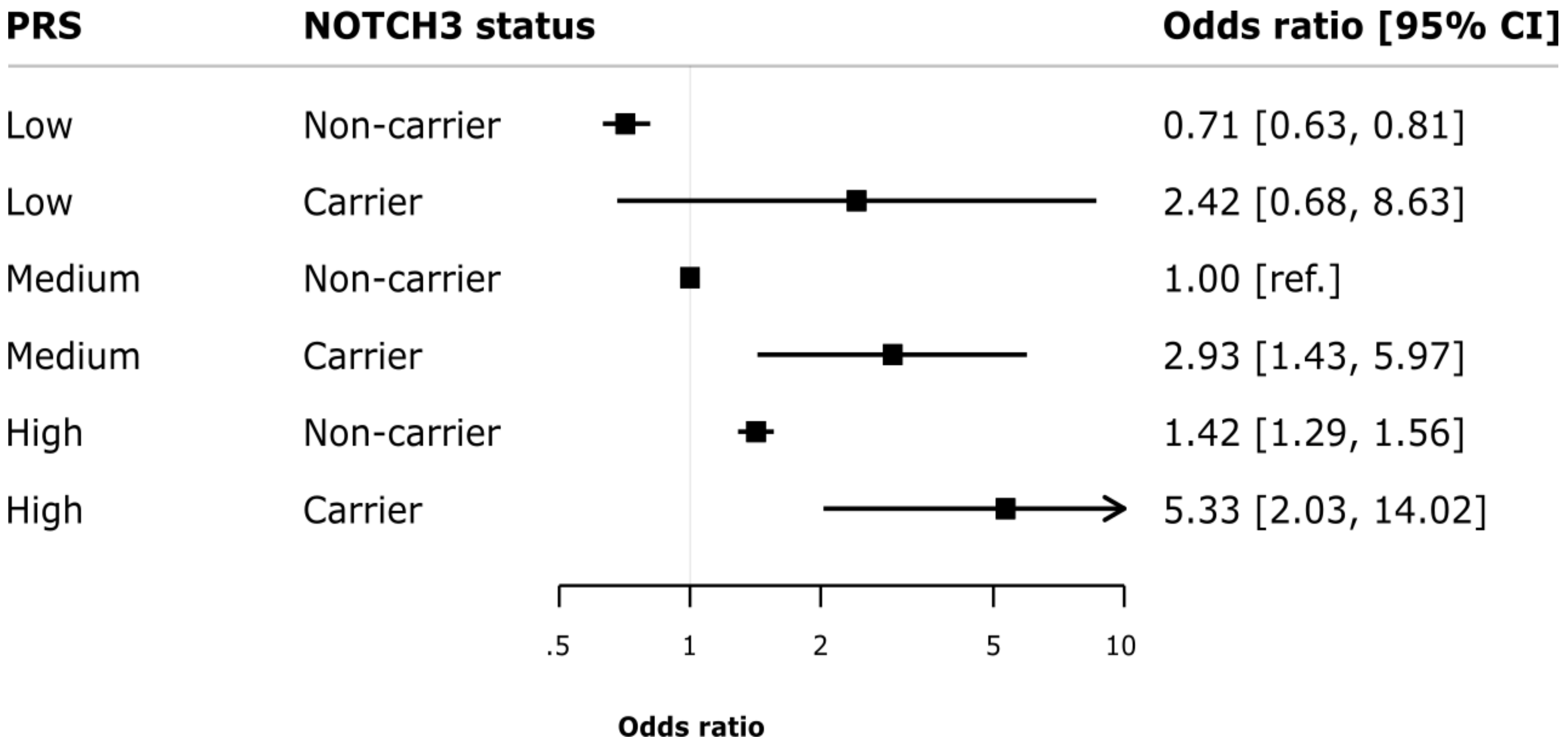


**Supplementary Figure 6. Forest plot displaying the odds ratios and associated 95% confidence intervals for ischaemic stroke stratified by polygenic risk score and *NOTCH3* status.**

P-value for a multiplicative continuous interaction with standardised score = 0.83; p-value for a multiplicative interaction with the categorical low, intermediate and high PRS groupings = 0.95; p-value for the additive interaction between continuous PRS assessed using the relative excess risk due to interaction (RERI) = 0.46.
